# Trends in Intervention Modality for Hospitalizations with Infectious Intracranial Aneurysms: A Nationwide Analysis

**DOI:** 10.1101/2024.09.27.24314522

**Authors:** Giana Dawod, Cenai Zhang, Hooman Kamel, Santosh Murthy, Neal S. Parikh, Alexander E. Merkler

**Affiliations:** Clinical and Translational Neuroscience Unit, Department of Neurology and Feil Family Brain and Mind Research Institute, Weill Cornell Medicine, New York, NY, USA

## Abstract

**Background/Objective:** Data regarding treatment of infectious intracranial aneurysms most effectively remains sparse. With growing utilization of endovascular therapy for cerebrovascular disease, we examined trends in endovascular versus neurosurgical treatment of infectious aneurysms and investigated the impact of treatment modality on outcomes.

**Methods:** Using data from the National Inpatient Sample from 2000 to 2019, we conducted a trends analysis on rates of treatment modalities among hospitalizations with infective endocarditis with ruptured or unruptured cerebral aneurysms. Treatment modalities were categorized as endovascular versus open neurosurgical repair based on ICD-9 and ICD-10 codes. Logistic regressions were utilized to assess the association between treatment modality and the outcomes of in-hospital mortality and discharge disposition.

**Results:** We identified 24,461 hospitalizations with an infectious intracranial aneurysm in the setting of infective endocarditis. Mean age was 56.0 years (SD, 17.8) and 61.8% were male. The overall rate of intervention was 5.8% (95% CI, 5.0-6.5%), and this did not change over time (p=0.669). There was a significant increase in the rate of endovascular repair (APC=3.6%; 95% CI, 1.2%-8.1%) and a significant decrease in the rate of open neurosurgical repair (APC= −5.4%; 95% CI, −8.1% to −3.5%). Treatment modality was not associated with in-hospital mortality (p=0.698) or non-home discharge disposition (p=0.897).

**Conclusion:** Although rates of infectious intracranial aneurysm intervention for infective endocarditis did not change, utilization of endovascular treatment increased while the use of open neurosurgical treatment decreased. Further directions include elucidating predictors of favorable outcomes for undergoing intervention and the most beneficial timing for the procedure during hospitalization.

**Summary:** *What is already known on this topic:* **-** Rates of infective endocarditis and infectious intracranial aneurysms continue to rise, however, there lacks a standard of care in management of this complication. Prior studies have only looked at national trends up until 2011, whereas ours not only analyzes trends up until 2019, but analyzes open neurosurgical and endovascular approaches separately.

*What this study adds:* - Our study indicates a significant increase in the use of endovascular treatment with a concomitant significant decrease in open neurosurgical clipping of infectious intracranial aneurysms. While patients undergoing any intervention had better mortality rates than patients treated with medical management alone, we found no statistical difference in mortality rates or disposition between the two treatment modalities.

*How this study might affect research, practice or policy:* - Our study highlights the need for further investigation of prognostic factors and timing of intervention in patients with infectious intracranial aneurysms, to standardize management to improve outcomes.

## Introduction

Infectious intracranial aneurysms (IIA) are among the extracardiac sequelae of infective endocarditis.^1^ Hematogenous spread of microbes to the cerebral vasculature, or direct extension of infection from the meninges, leads to vessel wall weakening and aneurysm formation.^1 2^ Once ruptured, the mortality rate may be as high as 80%.^3^ These aneurysms can be treated with antimicrobials, coiling, embolization, clipping, or a combination of these measures.^4^ Technical advancements such as smaller catheters and coils, coupled with increasing comfort of operators with these techniques, have made endovascular treatment of IIA increasingly feasible.^3 4^ While the proportion of patients with infective endocarditis with IIA undergoing any intervention has been stable over the past several decades,^5^ whether treatment approaches have shifted over time remains unclear. Further, the impact of treatment modality on patient outcomes is also not well understood. Therefore, we analyzed trends in treatment modality, specifically comparing trends in endovascular versus open neurosurgical treatment, in patients with infective endocarditis in the United States. We also assessed the association between intervention modality and in-hospital mortality.

## Methods

### Design

We performed a trends analysis and retrospective cohort studying using inpatient discharge data from the 2000-2019 releases of the National Inpatient Sample (NIS). The NIS is a de-identified, all-payer database that includes data for a representative sample of hospitalizations to non-federal hospitals in the United States, funded by the Agency for Healthcare Research and Quality.^6^ NIS data are released to researchers upon application, and we will make our analytical code available upon reasonable request. Analyses of NIS data were approved by the Weill Cornell Medicine institutional review board, and the need for informed patient consent was waived.

### Patient Population

We included all hospitalizations with infective endocarditis alongside a diagnosis of a possible ruptured cerebral aneurysm (on the basis of subarachnoid hemorrhage and intracerebral hemorrhage) or unruptured cerebral aneurysm. Diagnoses were ascertained using International Classification of Diseases, Ninth Revision, Clinical Modification (*ICD-9-CM)* codes. The tenth revision of the ICD coding was implemented in the U.S. on October 1, 2015; thus, after this point, the use of ICD-10-CM and PCS (Procedure Coding System) was implemented. The ICD-9-CM codes for infective endocarditis (421.0, 421.1, 421.9, 424.9, 424.90, 424.91, 424.99) and the ICD-10-CM equivalents (I33.0, I33.9, I38) have been used in prior studies, and have high sensitivity and positive predictive value for the diagnosis of infective endocarditis.^7–13^ Ruptured IIA were defined as the presence of subarachnoid hemorrhage (ICD-9-CM code 430; ICD-10-CM code I60) or intracerebral hemorrhage (ICD-9-CM codes 431; ICD-10-CM code I61) since both types of brain hemorrhages commonly occur in hospitalizations with IIA.^14–18^ Hospitalizations that had diagnoses of unruptured aneurysms alongside subarachnoid or intracerebral hemorrhage were also categorized as having ruptured IIA. Hospitalizations with unruptured aneurysms were identified using ICD-9-CM code 437.3 and ICD-10-CM code I67.1.^15^

### Measurements

We categorized hospitalizations as having undergone endovascular (ICD-9-PCS codes 39.52, 39.72, 39. 75, 39.76, 39.79; ICD-10-PCS codes 03VG3BZ, 03VG3CZ, 03VG3DZ, 03VG3HZ,03VG3ZZ, 03LG3BZ, 03LG3CZ, 03LG3DZ, 03LG3ZZ) versus open surgical (ICD-9-PCS code 39.51, 38.81; ICD-10-PCS codes 03VG0BZ, 03VG0CZ, 03VG0DZ, 03VG0HZ, 03VG0ZZ, 03LG0BZ, 03LG0CZ, 03LG0DZ, 03LG0ZZ) IIA treatment using ICD procedure codes.^5 16–19^ We tabulated several covariates to describe our study population. The NIS includes demographic variables such as age, sex, race, comorbidities, and insurance status. In terms of patient outcomes data, we focused on in-hospital mortality and discharge disposition. For discharge disposition, a favorable outcome was defined as a routine discharge to home. Unfavorable outcomes encompassed discharge to an acute rehabilitation hospital, a skilled nursing facility, hospice, or in-hospital death. Any hospitalizations lacking or with unclear discharge disposition data were excluded from the analysis.

### Statistical Analysis

First, we evaluated trends in the rates of endovascular and open surgical treatments of IIA over time. In the primary approach, we analyzed these trends among hospitalizations with both unruptured and ruptured IIA. In secondary analyses, we focused on trends among people with a ruptured IIA defined as subarachnoid or intracerebral hemorrhage, and then specifically as subarachnoid hemorrhage alone. All rates were age-standardized using US Census data for the trend analysis. We employed a log-linear model in Joinpoint Software, which is designed for trends analyses using joinpoint models where multiple trends can be identified and connected at inflection points or “joinpoints”, when applicable.

Second, among hospitalizations with IIA treatment, logistic regression was utilized to assess the association between treatment modality (open versus endovascular) and outcomes, including in-hospital mortality and discharge disposition. The regression analysis adjusted for age, sex, race/ethnicity, insurance type, rupture status, and the presence of comorbidities including hypertension, diabetes, atrial fibrillation, coronary heart disease, congestive heart failure, peripheral vascular disease, chronic kidney disease, valvular heart disease, tobacco use and alcohol abuse (Table 1). Hospitalizations involving both endovascular and open surgical repair were excluded from the analysis due to the low sample size, which was insufficient to estimate the national metrics accurately.

**Table 1:** Characteristics of Patients Hospitalized with Cerebral Aneurysm in the Setting of Infective Endocarditis from 2000 to 2019, Stratified by Treatment.

|  | No Intervention | Endovascular treatment | Neurosurgical treatment | Both |
| --- | --- | --- | --- | --- |
| **Number of Patients** | 23,054 | 388 | 979 | 40 |
| **Age, Years, mean (SD)** | 57 (18) | 45 (18) | 48 (16) | 44 (17) |
| **Female** | 8826 (38%) | 126 (32%) | 373 (38%) | 20 (50%) |
| **Race/ethnicity** |  |  |  |  |
| Non-Hispanic White | 15,570 (68%) | 291 (75%) | 706 (72%) | 30 (75%) |
| Non-Hispanic Black | 3,624 (16%) | 34 (9%) | 126 (13%) | 10 (25%) |
| Hispanic | 2,237 (10%) | 39 (10%) | 75 (8%) | - |
| Asian or Hawaiian or Native American | 722 (3%) | 14 (4%) | 29 (3%) | - |
| Other | 901 (4%) | 10 (2%) | 44 (4%) | - |
| **CCI, median (IQR)** | 2 (1-4) | 1 (1-2) | 2 (1-3) | 1 (1-2) |
| Abbreviations: SD, standard deviation; SAH, subarachnoid hemorrhage; ICH, intracranial hemorrhage; CCI, Charlson Comorbidity Index; |  |  |  |  |

To account for the NIS sample redesign, we used the updated trend weights for the period between 2000 and 2011 and the original discharge weights for the years 2012 to 2019. We applied sampling weights provided by the NIS to obtain national estimates for all outcomes. All trend analyses were two-tailed with statistical significance defined as a P value <0.05. Analyses were performed using Stata (version 15.1, College Station, Texas) and Joinpoint (version 5.1.0, National Cancer Institute).

## Results

We identified 24,461 hospitalizations with a ruptured or unruptured cerebral aneurysm in the setting of infective endocarditis, of which 22,685 (92.7%) were hospitalizations with a ruptured IIA and 1,776 with a unruptured aneurysm. The mean age of hospitalizations was 56.0 years (SD, 17.8), with males comprising 61.8% of the total hospitalizations.

Overall, 1,407 hospitalizations underwent IIA treatment. Among them, 428 received open neurosurgical repair, 1,019 received endovascular repair, and 40 underwent both types of repairs. The overall rate of IIA intervention over the entire study period was 5.8% (95% CI, 5.0%-6.5%) and this rate did not change appreciably over time (p=0.669). The age-standardized rate of endovascular repair increased from 0% (95% CI, −0.1% to 0.1%) in 2000 to 3.2% (95% CI, 1.6% to 4.8%) in 2019. This increase was statistically significant, with an annual percent change (APC) of 3.6% (95% CI, 1.2%-8.1%; p=0.007) over the studied period. On the other hand, there was a notable decrease in the rate of open neurosurgical repair during the same period. The age-standardized rate of open neurosurgical repair declined from 3.8% (95% CI, 2.1% to 5.6%) in 2000 to 0.6% (95% CI, −0.1% to 1.2%) in 2019. This decline was statistically significant, with an annual percent change (APC) of −5.4% (95% CI, −8.1% to −3.5%; p<0.001) over the studied period (Figure 1). Trends were similar when restricting to hospitalizations with either ruptured IIA or hospitalizations exclusively with SAH (Tables 2 and 3).

**Figure 1.**
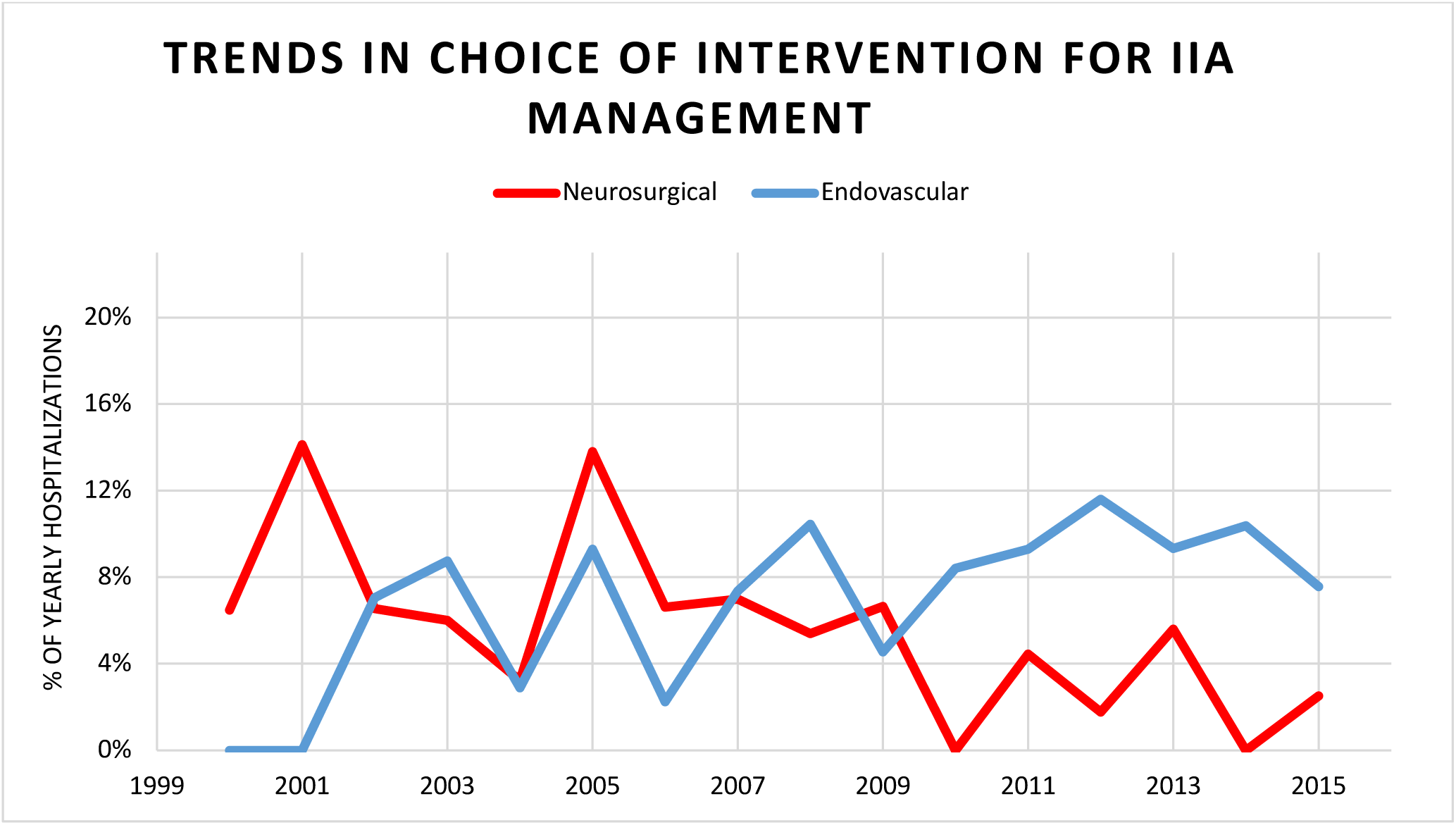
National Trends in Intervention Modality for IIA Management, 2000-2019.

**Table 2:** Hospitalization Metrics Among Patients with IIA Who Underwent either Neurosurgical or Endovascular Intervention.

|  | **Endovascular Therapy** | **Neurosurgical Treatment** |
| --- | --- | --- |
| **In-hospital Mortality (%)** | 16.0% (10.7%-21.3%) | 19.8% (10.9%-28.8%) |
| **Non-home Discharge (%)** | 82.4% (77.1%-87.8%) | 86.4% (78.9%-93.9%) |

**Table 3.** Trends in Endovascular and Open Neurosurgical Repair of Presumed IIA, from 2000 to 2019.

|  | **\*Trends in Endovascular Treatment** | **\*Trends in Neurosurgical Treatment** |
| --- | --- | --- |
| **All MA** | 3.6% (1.2%-8.1%) | -5.4% (-8.1% to -3.5%) |
| **Ruptured (ICH+SAH)** | 4.6% (1.6%-10.7%) | -4.7% (-8.8% to -1.9%) |
| **Ruptured (SAH only)** | 3.3% (1.0%-8.0%) | -6.8% (-12.9% to -2.3%) |
| *Data are presented as APC (Annual Percent Change) with 95% confidence intervals. |  |  |

The overall in-hospital mortality rate among hospitalization with infective endocarditis and IIA was 31.6% (95% CI, 30.2%-32.9%). Hospitalization with any IIA intervention had an in-hospital mortality rate of 17.3% (95% CI, 12.7%-22.0%), whereas the rate was 32.4% (95% CI, 31.0-33.8%) in hospitalizations without intervention. Among hospitalizations with any intervention, in-hospital mortality was lower in hospitalizations with endovascular (16.0%; 95% CI, 10.7% to 21.3%) versus open surgical (19.8%; 95% CI, 10.9% to 28.8%) repairs. The in-hospital mortality rate did not show a significant difference between hospitalizations with endovascular repair and open surgical repair (p=0.698). Moreover, there was also no significant difference in the in-hospital mortality rate in sensitivity analyses that were restricted to hospitalizations with ruptured intracranial aneurysms (p=0.630) or subarachnoid hemorrhage (p=0.459).

Among all hospitalizations with infective endocarditis and IIA, unfavorable discharge disposition occurred in 82.4% (95% CI, 77.1%-87.8%) of those with endovascular repair, compared to 86.4% (95% CI, 78.9%-93.9%) of those with open surgical repair (p=0.897). There was no significant difference in discharge disposition between endovascular and open surgical repair, nor in the sensitivity analysis restricted to hospitalizations with ruptured intracranial aneurysms (p=0.451) or subarachnoid hemorrhage (p=0.536).

## Discussion

Within a large, randomized national sample of hospitalizations across the U.S., we found that from 2000 to 2019, the rates of interventions for IIA remained relatively unchanged. Despite this finding however, the rates of endovascular treatment for IIA management increased at the same time as rates of open neurosurgical treatment decreased. Our study adds to the existing literature that attempts to analyze the trends in interventions. Specifically, we compared the types of intervention methods for IIA in IE patients. Earlier studies that looked at national trends found that although mortality rates were significantly higher in the patient groups that were treated conservatively, the proportion of patients receiving neurosurgical procedures, both neurosurgical clipping and endovascular coiling, remained constant throughout the years.^5^ With the advent of technological advancements and increased scope for which endovascular management is utilized in the last decade alone, we believe there to be a growing role for endovascular intervention in IIA treatment.

Given that 65% of patients with IIA are found to have underlying infective endocarditis, a multi-disciplinary approach is warranted for optimal outcomes.^1 4^ In the case of ruptured aneurysms, the mortality rate has proven to be higher for those conservatively treated compared to group that underwent intervention.^3^ Treatment with antibiotics alone in IE patients with IIA comes with its own set of risks including possible antibiotic resistance due to biofilm formation and enlargement of the aneurysm.^1^ In several studies assessing outcomes of conservative treatment in unruptured aneurysms, more than 20% of IIA continued to grow despite appropriate antibiotic use.^20^ The choice to intervene must take into consideration the low risk of rupture against the high mortality rate once ruptured.^21^ Ultimately, the complexity of IIA management has been amplified by several considerations prior to intervention. These include the patient’s clinical stability, recent cardiothoracic surgery to address the underlying etiology of the IE, patient’s age, and anatomical characteristics of the aneurysm itself, especially if it is unruptured.^5^

The decline in cases in which neurosurgical clipping is utilized coincides with adoption of newer techniques and tools to address IIA in both a more timely and safe manner. Additionally, the rapid growth of the elderly population, an already high-risk surgical cohort, is accompanied by an increase in cardiac disease including degenerative valvular disease, increasing susceptibility to IE.^22^ Neurosurgical clipping of IIA poses an inherent risk with the morphological challenge of successful obliteration of the aneurysm including little to no aneurysmal neck, increased vessel friability, and surrounding vessel involvement.^23^ In the context of left-sided heart disease, patients with paravalvular abscesses, recurrent systemic embolization, confirmed heart failure, valve dysfunction, large mobile vegetations, and persistent infection despite an appropriate duration of antibiotic therapy require cardiothoracic intervention.^22^ Depending on institution and status of patient, this may occur prior to securing the aneurysm, especially in light of studies showing there to be mortality benefit in complicated IE patients undergoing valve surgery earlier.^22 24^ Data looking at the temporal relationship between cardiac and neurological intervention in IE patients show that in the cases where aneurysm management preceded valve surgery, more than half of the interventions were completed endovascularly.^24^ This may be due to operators preferring to delay neurosurgical clipping in order to allow for the inflammatory process to subside, giving a chance for the aneurysm to become fibrotic with reduced likelihood to rupture when being operated on.^25^

In the case of proximal IIA, the use of stent assisted coiling and flow diversion therapy have proven to be successful for embolization.^20^ For more distal aneurysms, liquid embolic agents (LEA) such as N-butyl cyanoacrylate (NBCA) and ethyl vinyl alcohol copolymer in dimethyl sulfoxide solution, also known as Onyx, are used in addition to parent artery occlusion. Initial apprehension with regards to endovascular treatment stemmed from the concept of deploying devices into a bacteremic patient, posing the risk of possible abscess formation. However, this rare complication has only been documented in older case studies at the onset of endovascular therapy utilization.^3 26^ The advantages of these LEAs include the reduced potential for infection, no required antiplatelet loads or maintenance antiplatelet use, and the ability to control the delivery of the agent into the smallest vessels.^27^ Onyx remains more utilized given that it is non-adhesive and is visible under fluoroscopy for more controlled obliteration of the aneurysm. Additionally, the studies utilizing LEAs intracranially do not report major strokes or hemorrhagic transformations distal to the vascular territory that were embolized.^21 28^

Despite the dynamic progress made in the management of IIA in the last decade, further analysis in the imaging protocols in institutions for IE patients both with and without neurological symptoms may be warranted. It is important to know if the rate of detection of the aneurysms itself is impacting the rate of intervention. Although serial imaging and conservative therapy has been recommended in the past for unruptured aneurysms, the rapid rate in which IIA can grow may warrant more aggressive screening.^29 30^

The results of our trends analysis should be taken in the context of the inherent limitations of this study. This was a retrospective analysis of the NIS database, leaving room for selection bias in our patient cohort. Information regarding the temporal relationships in the patients’ hospital course is difficult to be derived in the database and depends on other factors not accounted for. Specific codes for ruptured and unruptured IIA do not exist. While we had utilized previous validated ICD coding to help in delineating these groups, our own definitions of these two separate entities for analysis likely introduced variability and may not have captured all hospitalizations of interest. The ICD coding for SAH as well can be synonymous with ruptured intracranial aneurysms. A SAH or ICH along with the diagnosis of IE is what was defined as ruptured IIA. Additionally, our analysis was limited by the available variables including lack of knowledge of the infectious organism, the characteristics and severity of the valvular disease, and lack of information regarding the ultimate cause of mortality.

## Conclusions

This study demonstrates a national trend towards the utilization of endovascular treatment for IIA in patients with underlying infective endocarditis despite an overall consistent rate in the use of intervention from 2000-2019. Patients undergoing both neurosurgical and endovascular intervention versus medical management alone demonstrated a better mortality rate, however, the significance of this phenomenon is unclear. While there was no clear benefit in hospital disposition or mortality outcomes, further investigation is warranted to understand the importance of timing of the intervention relative to the diagnosis of IIA. Additionally, further analyses should delineate prognostic factors to determine the candidates most likely to benefit from intervention versus medical management alone.

## Data Availability

Available upon requestn

